# Acute Cardiovascular Effects of Psilocybin: A Pooled Analysis of 14 Studies with Safety Recommendations

**DOI:** 10.64898/2026.04.28.26351625

**Authors:** Sandeep M. Nayak, Nathan D. Sepeda, Matthew Nielsen Dick, Praachi Tiwari, Zarmeen Zahid, Ceyda Sayalı, Brandon M. Weiss, David B. Yaden, Albert Garcia-Romeu, Brian S. Barnett, Frederick S. Barrett

## Abstract

**Background:** Psilocybin has increasingly been studied as a therapeutic for psychiatric and neurologic conditions, yet comprehensive cardiovascular safety data are limited. Current trials often exclude individuals with blood pressure ≥140/90 mmHg, criteria established conservatively without robust empirical support.

**Objective:** Characterize the blood pressure and heart rate response to typical therapeutic doses of psilocybin and provide an evidence base for cardiovascular eligibility criteria and monitoring protocols for future clinical trials and emerging therapeutic practice.

**Methods:** We pooled data from 536 psilocybin sessions (oral doses 20–47 mg) among 368 participants across 14 studies at Johns Hopkins University since 1999. Blood pressure and heart rate were measured at baseline and at least hourly up to 360 minutes post-administration. We quantified peak changes, threshold excursions, and excursion duration.

**Results:** Psilocybin produced modest, transient blood pressure elevations. Median peak systolic blood pressure (SBP) was 145 mmHg (IQR 134–156), representing a median increase of 22 mmHg from baseline. Blood pressure peaked at approximately 90 minutes and returned to near-baseline by 300 minutes. SBP exceeded 170 mmHg in 32 sessions (6.0%; median duration 8.5 minutes) and 180 mmHg in 17 sessions (3.2%; median duration 10 minutes). Antihypertensive medication was administered in only 1 session (0.2%). Higher baseline blood pressure was associated with smaller increases, suggesting a ceiling effect rather than exaggerated response.

**Conclusions:** Psilocybin produces modest, transient blood pressure elevations comparable to moderate exercise. Current exclusion criteria of ≥140/90 mmHg are not supported by these data. We recommend broadening eligibility to <160/100 mmHg while maintaining exclusions for established cardiovascular disease.

## Introduction

Psilocybin, a serotonergic psychedelic, has demonstrated promising therapeutic potential across several psychiatric conditions. It is being actively investigated for a range of indications including depression (Davis et al., 2021; Goodwin et al., 2022; Raison et al., 2023), substance use disorders (Bogenschutz et al., 2022; Johnson et al., 2026; Rieser et al., 2025), and post-traumatic stress disorder (PTSD) (McGowan et al., 2025), among others. As psilocybin advances toward possible regulatory approval, establishing comprehensive safety parameters for administration is increasingly important.

Psilocybin is known to produce transient elevations in blood pressure and heart rate (Hasler et al., 2004; Nahlawi et al., 2025; Wsoł, 2023). The mechanism is multifactorial, involving direct serotonin 2A (5-HT_2A_) receptor-mediated vasoconstriction and sympathomimetic effects related to both receptor interactions and psychological arousal. Peripheral arterial vasoconstriction by serotonergic agents appears to require concomitant activation of both 5-HT_2A_ and 5-HT_1B_ receptors; notably, psilocybin has relatively low affinity for the 5-HT_1B_ receptor compared to other psychedelics such as LSD, which may limit its vasoconstrictive potential (Nahlawi et al., 2025; Wsoł, 2023). Consequently, exclusion of individuals with elevated blood pressure has been a longstanding practice in clinical trials with psychedelics (Johnson et al., 2008).

However, these cardiovascular exclusion criteria lack a robust empirical basis specific to psilocybin administration, and recent reviews have highlighted cardiovascular safety as a critical knowledge gap in the field (Nahlawi et al., 2025). Overly conservative thresholds may unnecessarily exclude patients who could safely benefit from psilocybin therapy, particularly those with hypertension—a population that substantially overlaps with individuals suffering from depression and substance use disorders among many other disorders that are currently receiving attention as potential therapeutic targets of psilocybin.

To address this critical evidence gap, we conducted a pooled analysis of psilocybin dosing sessions from the Johns Hopkins Center for Psychedelic and Consciousness Research, the world’s largest academic psychedelic research program. This dataset of 536 monitored sessions across diverse clinical populations and indications represents the most comprehensive characterization of acute cardiovascular responses to psilocybin to date. Our aims were to quantify the magnitude and temporal dynamics of blood pressure elevation from baseline in these sessions and provide an empirical foundation to inform evidence-based eligibility criteria and safety monitoring protocols for both clinical trials and emerging therapeutic practice.

## Methods

### Study Design and Data Sources

We conducted a pooled analysis of cardiovascular safety data from psilocybin administration sessions across clinical studies conducted at Johns Hopkins University School of Medicine. Studies included trials investigating psilocybin for cancer-related distress (Griffiths et al., 2016), major depressive disorder (Davis et al., 2021), anorexia nervosa (NCT04052568), PTSD (NCT06407635), Alzheimer’s disease-related distress (NCT04123314), Lyme disease-related symptoms (Garcia-Romeu et al., 2026), and Obsessive-Compulsive Disorder (OCD; NCT05546658), as well as studies in healthy volunteers examining psilocybin’s subjective effects (Griffiths et al., 2006), dose-response (Griffiths et al., 2011), effects on spiritual practice (Griffiths et al., 2018), effects in religious professionals (Griffiths et al., 2025), effects in long-term meditators (NCT01988311), comparison with dextromethorphan (DXM) (Barrett et al., 2018), and a functional magnetic resonance imaging (fMRI) study (Barrett et al., 2020). With the exception of ongoing blinded trials, this dataset represents all psilocybin studies conducted at Johns Hopkins University for which the Principal Investigator provided permission to include data. All studies were approved by a Johns Hopkins Medicine Institutional Review Board, and all participants provided written informed consent.

### Participants

Participants were adults meeting a range of different study-specific eligibility criteria, described in detail in the primary publications and/or on clinicaltrials.gov. Inclusion criteria for blood pressure varied across studies, ranging from <140/90 mmHg to <165/95 mmHg. All studies excluded individuals with major cardiovascular conditions including arrhythmias, prolonged QT interval, stroke, coronary artery disease, etc.

### Psilocybin Administration

Psilocybin was administered orally in capsule form. We included only sessions with doses ≥20 mg, as lower doses are generally not representative of therapeutic administrations. Some studies administered bodyweight-adjusted doses (e.g., 20 mg/70 kg), while others administered fixed absolute doses, which has generally become standard practice in clinical trials (Garcia-Romeu et al., 2021). Sessions were conducted in a controlled research environment with trained facilitators present throughout, and participants were monitored for 6–8 hours following administration.

### Cardiovascular Monitoring

On dosing days, sessions proceeded if pre-dose blood pressure was below study-specific thresholds (typically <140/90 to <155/95 mmHg).

Blood pressure (systolic and diastolic) and heart rate were measured at baseline (pre-administration) and at scheduled intervals post-administration (30, 60, 90, 120, 180, 240, 300, and 360 minutes). Elevated readings triggered increased monitoring frequency (every 5 minutes), with thresholds varying slightly across studies but generally systolic blood pressure (SBP) >170 mmHg, diastolic blood pressure (DBP) >100–110 mmHg, or HR >110–120 beats per minute (bpm). Antihypertensive medications (including sublingual nitroglycerin, or oral clonidine, nifedipine, or captopril, depending on study protocol) were available to treat sustained elevations at physician discretion, though some studies protocolized antihypertensive administration (e.g. administer if blood pressure remained above 200 SBP and/or 110 DBP for 15 minutes).

### Outcomes

The primary outcomes examined in the present analysis were:

1. Peak systolic and diastolic blood pressure during each session
2. Peak change from baseline in SBP and DBP
3. Proportion of sessions exceeding the following blood pressure thresholds (SBP >170, >180 mmHg, >200 mmHg; DBP >100, >110 mmHg; severe hypertension defined per ACC/AHA guidelines as SBP/DBP >180/120 (Jones et al., 2025))
4. Duration of threshold excursions

Secondary analyses examined the relationship between baseline blood pressure and peak blood pressure response, and peak heart rate response.

## Statistical Analysis

Blood pressure and heart rate timecourse were visualized using locally estimated scatterplot smoothing (LOESS) with 95% confidence intervals.

For each session, we identified peak systolic and diastolic blood pressure values and peak heart rate. Peak values and excursion durations are summarized as median (IQR) given right-skewed distributions, with 95th percentiles reported to characterize the upper range relevant to safety monitoring. Time to peak was defined as the median time post-dose at which peak values occurred. Baseline characteristics and mean change from baseline at specified timepoints are reported as mean (SD or 95% CI).

To inform duration of hemodynamic monitoring, we identified the timepoint at which blood pressure visually appeared to return to baseline on LOESS curves and calculated mean change from baseline with 95% confidence intervals at that timepoint.

We calculated the proportion of sessions exceeding predefined blood pressure thresholds (SBP >170, >180 mmHg; DBP >100, >110 mmHg) and heart rate thresholds (>100, >110, >120 bpm) with exact binomial 95% confidence intervals.

To evaluate the relationship between baseline cardiovascular parameters and hemodynamic response, we used linear regression with peak change from baseline as the dependent variable and baseline value as the predictor. We additionally fit multivariable linear regression models for peak SBP and DBP change, incorporating age, sex, and weight as covariates alongside baseline blood pressure, to evaluate independent predictors of hemodynamic response. For blood pressure, we also compared peak changes between sessions with baseline BP ≥140/90 mmHg versus <140/90 mmHg. For heart rate, we compared sessions with baseline HR ≥100 bpm versus <100 bpm.

In supplementary analyses, we used linear mixed-effects models with random intercepts for participant and study to assess whether dose or session number predicted peak blood pressure change (see Supplementary Materials).

Analyses were conducted in R version 4.4.1 (R Foundation for Statistical Computing, Vienna, Austria).

## Results

### Sample

We analyzed 536 psilocybin sessions among 368 participants across 14 studies. Among the 368 participants, mean age was 44.8 years (SD = 13.8; range 21–82; n = 365); 55.6% (n = 203/365) were female; and 89.1% (n = 295/331) were White. Psilocybin doses ranged from 20 to 47.4 mg. Mean baseline SBP was 121.3 mmHg (SD 12.4), DBP 71.6 mmHg (SD 9.2), and heart rate 70.1 bpm (SD 13.1).

**Table 1.** Summary of included studies.

| Study | Participants | Sessions | Dose range (mg) |
| --- | --- | --- | --- |
| Spiritual Practices (Griffiths et al., 2018) | 72 | 110 | 20.2-42.7 |
| Anorexia nervosa (NCT04052568) | 20 | 61 | 20-30 |
| OCD (NCT05546658) | 30 | 60 | 20-30 |
| PTSD (NCT06407635) | 21 | 38 | 25-35 |
| Major Depression (Davis et al., 2021) | 24 | 37 | 20-41.4 |
| Cancer distress (Griffiths et al., 2016) | 37 | 37 | 20-40.8 |
| Healthy volunteers, subjective effects (Griffiths et al., 2006) | 35 | 35 | 22.6-45.1 |
| Long-term meditators (NCT01988311) | 34 | 34 | 20.9-38.2 |
| Religious Professionals (Griffiths et al., 2025) | 18 | 31 | 20.4-43.5 |
| Dose effects (Griffiths et al., 2011) | 18 | 27 | 20.4-47.4 |
| Psilocybin vs DXM (Barrett et al., 2018) | 21 | 26 | 20.3-44.7 |
| Lyme disease (Garcia-Romeu et al., 2026) | 18 | 18 | 25 |
| Alzheimer's distress (NCT04123314) | 10 | 12 | 20.1-33.5 |
| Psilocybin fMRI (Barrett et al., 2020) | 10 | 10 | 20.2-32.6 |

### Blood Pressure and Heart Rate Timecourse

Blood pressure peaked at approximately 90 minutes post-dose and declined steadily thereafter. Visual inspection of LOESS curves (Figure 1A) indicated blood pressure returned to near-baseline by 300 minutes (5 hours); mean change from baseline at this timepoint was +2.8 mmHg systolic (95% CI 1.7 to 3.8) and +0.7 mmHg diastolic (95% CI -0.1 to 1.5).

**Figure 1.**
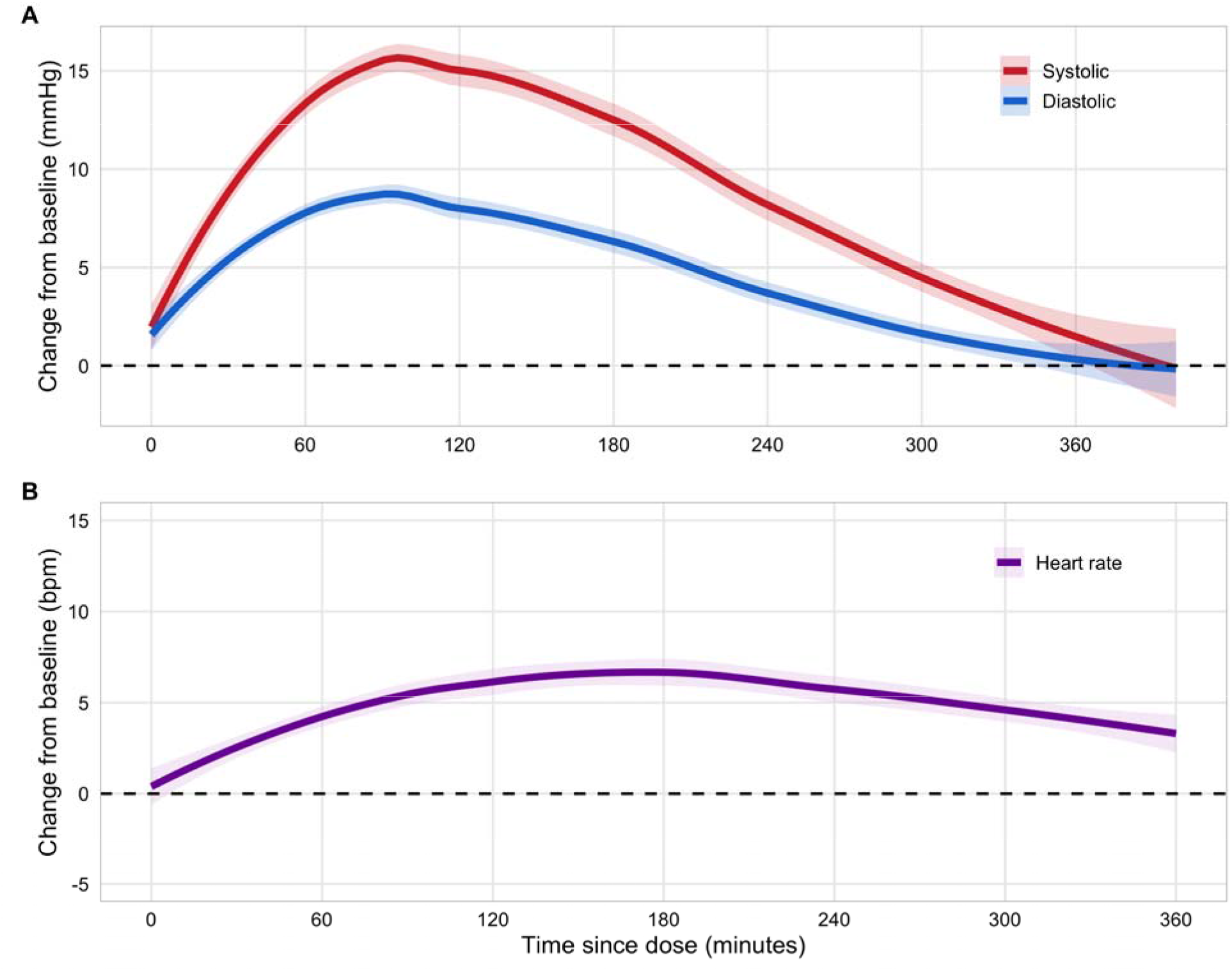
Timecourse of mean cardiovascular changes following psilocybin ≥20mg. (A) Blood pressure (n = 536 sessions). (B) Heart rate (n = 534 sessions). LOESS smooth with 95% confidence intervals.

**Figure 2.**
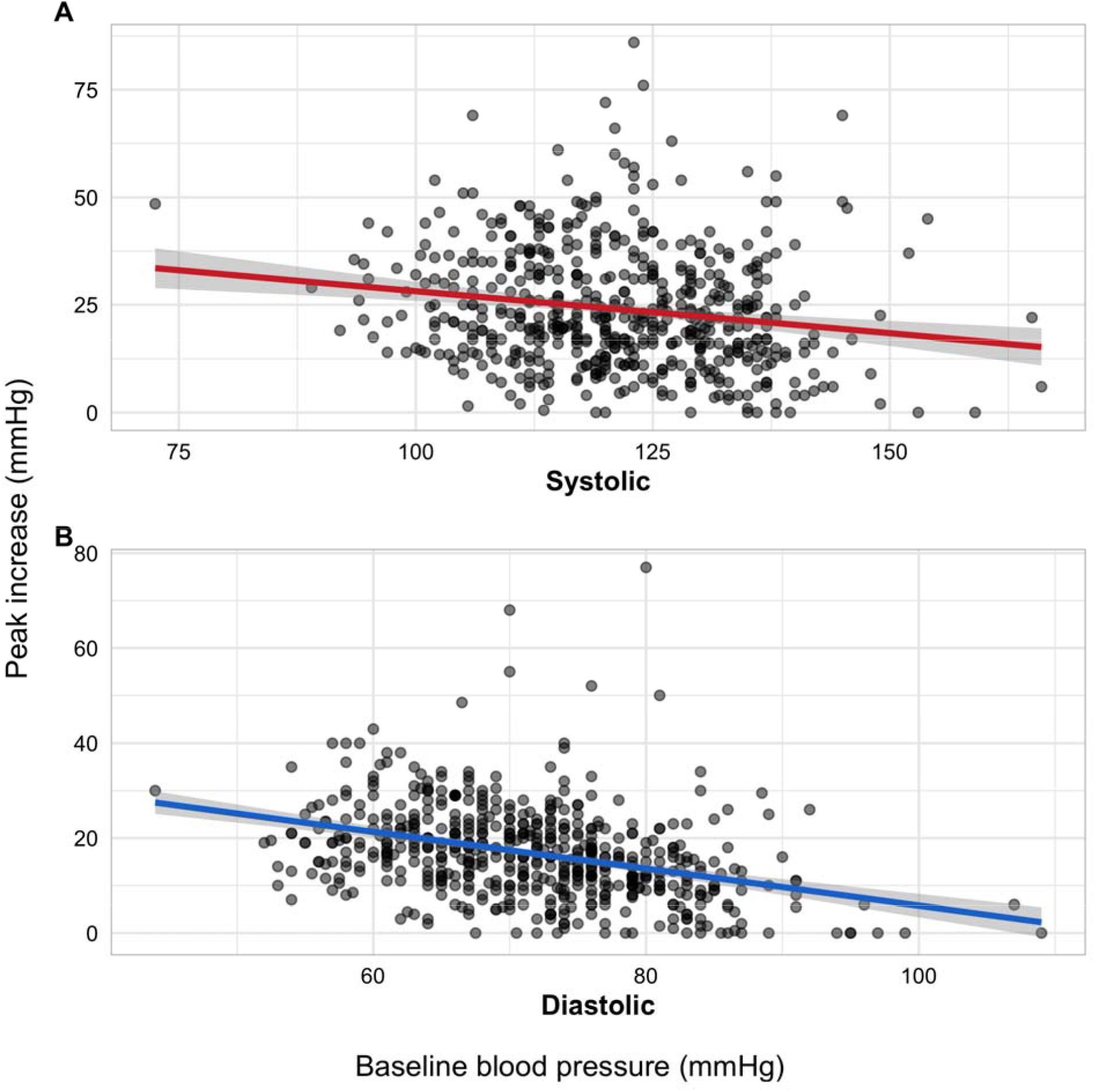
Association between baseline blood pressure and peak blood pressure increase. (A) Peak systolic blood pressure (β = -2 mmHg per 10 mmHg, p < 0.001). (B) Peak diastolic blood pressure (β = -3.9 mmHg per 10 mmHg, p < 0.001). Lines represent linear regression with 95% confidence intervals.

Heart rate peaked later, at approximately 180 minutes post-dose, and remained modestly elevated at 300 minutes (Figure 1B); mean change from baseline at this timepoint was +3.2 bpm (95% CI 2.1 to 4.2). Heart rate data was unavailable for post-baseline timepoints in one session and baseline heart rate was unrecorded in a second session; both sessions were excluded from heart rate analyses.

### Peak Blood Pressure and Heart Rate

Median peak SBP was 145 mmHg (IQR 134–156; 95th percentile 174 mmHg). Median peak DBP was 89 mmHg (IQR 82–95; 95th percentile 103 mmHg). Median peak increase from baseline was 22 mmHg systolic (IQR 14–33; 95th percentile 49 mmHg) and 16 mmHg diastolic (IQR 10–22; 95th percentile 33 mmHg). The mean change at any given time point (Figure 1A) was lower (∼15 mmHg systolic at maximum) because individual peaks occurred at different times and were averaged with non-peak values.

Median peak heart rate was 85 bpm (IQR 74–95; 95th percentile 115 bpm), representing a median increase of 14 bpm from baseline (IQR 7–23; 95th percentile 37 bpm).

### Threshold Excursions

Significant blood pressure elevations were infrequent and brief. SBP exceeded 170 mmHg in 32 sessions (6% [95% CI 4.1–8.3]; median duration 8.5 min [IQR 5–28]), 180 mmHg in 17 sessions (3.2% [95% CI 1.9–5.0]; median duration 10 min [IQR 5–48]), and 200 mmHg in 2 sessions (0.4% [95% CI 0.0–1.3]; median duration 10 min [IQR 8–10]). DBP exceeded 100 mmHg in 40 sessions (7.5% [95% CI 5.4–10.0]; median duration 8 min [IQR 5–30]) and 110 mmHg in 15 sessions (2.8% [95% CI 1.6–4.6]; median duration 15 min [IQR 5–50]).

2 sessions (0.4% [95% CI 0.0–1.3]) met criteria for ACC/AHA defined severe hypertension (SBP >180 mmHg and DBP >120 mmHg simultaneously).

Only 1 session (0.2%) involved administration of antihypertensive medication. This occurred in the cancer distress trial (Griffiths et al., 2016); a 50s male participant who received 29 mg psilocybin developed asymptomatic SBP of 214 mmHg (baseline blood pressure 145/84), was given nitroglycerin per protocol, and was discharged at the usual timepoint without further issues. No participants experienced adverse sequelae or symptoms as a result of their transient elevations in blood pressure.

Heart rate exceeded 100 bpm in 89 sessions (16.6%), 110 bpm in 39 sessions (7.3%), and 120 bpm in 17 sessions (3.2%); all elevations were self-limited and required no intervention.

### Baseline Blood Pressure and Heart Rate and Response

Higher baseline blood pressure was associated with smaller blood pressure increases during psilocybin sessions. For each 10 mmHg higher baseline SBP, peak SBP increase was 2.0 mmHg smaller (β = -0.20, p < 0.001). This relationship was stronger for DBP (β = -0.39, p < 0.001). Participants had baseline BP ≥140/90 mmHg in 34 sessions (6.1%). These participants showed smaller increases from baseline compared to those with BP <140/90 at baseline (median ΔSBP 15 vs 22 mmHg; median ΔDBP 11 vs 16 mmHg), suggesting a ceiling effect rather than an exaggerated response.

In multivariable analysis adjusting for age, sex, and weight, ceiling effects persisted — participants with higher baseline BP showed attenuated peak increases (SBP: β = -0.31 per mmHg, p < 0.001; DBP: β = -0.40 per mmHg, p < 0.001). Older participants showed larger peak blood pressure increases, with each additional decade of age associated with approximately 4 mmHg greater peak SBP rise (β = 0.39 per year, p < 0.001) and 1 mmHg greater peak DBP rise (β = 0.10 per year, p < 0.001). Higher body weight was associated with modestly larger SBP increases (β = 0.10 mmHg per kg, p = 0.018). Sex was not significantly associated with BP response.

A similar pattern was observed for heart rate. For each 10 bpm higher baseline HR, peak HR increase was 1.3 bpm smaller (β = -0.13, p = 0.001). Participants had baseline HR ≥100 bpm in 11 sessions (2%). These participants showed smaller increases from baseline than those with baseline HR <100 bpm (median ΔHR 5 vs 14 bpm).

In mixed-effects models, dose was not significantly associated with peak SBP change (β = 0.04 mmHg per mg, p = 0.39) nor peak DBP change (β = 0.07 mmHg per mg, p = 0.08); session number did not predict BP response; very little variance was accounted for between studies (Supplementary Tables 1 and 2).

## Discussion

This pooled analysis of 536 high-dose psilocybin sessions represents the largest characterization of psilocybin’s acute cardiovascular effects to date.

The hemodynamic profile identified is one of modest, transient, and self-limited blood pressure elevation. Peak blood pressure elevations were comparable to those observed during moderate physical activity (Fletcher et al., 2013) and sex (Levine et al., 2012), both of which are considered safe for patients with stable cardiovascular disease. The time-course of blood pressure elevation we observed is consistent with psilocybin’s pharmacology, with blood pressure peaking at approximately 90 minutes, corresponding to peak psilocin plasma concentrations. Heart rate peaked later, at approximately 180 minutes, possibly reflecting sustained autonomic arousal beyond psilocin’s peak pharmacokinetic window. Blood pressure returned to near-baseline by 5 hours, providing a clear timeframe for hemodynamic monitoring. These kinetics are predictable and reproducible across 14 heterogeneous studies and multiple clinical indications including individuals from 21 to 82 years of age. Despite 57 sessions (10.6%) with participants exceeding SBP >170 or DBP >100 mmHg, an antihypertensive was administered in only 1 session (0.2%), and no clinically significant sequelae were reported, further supporting the self-limiting nature of these elevations.

Contrary to concerns that elevated baseline blood pressure might produce exaggerated responses, we observed the opposite. Higher baseline BP was associated with attenuated peak increases, suggesting a ceiling effect. Participants with baseline BP ≥140/90 mmHg showed smaller increases from baseline; their higher absolute peak values simply reflect their starting point. This is a notable difference from ketamine, where patients with baseline hypertension show greater maximal changes in both systolic and diastolic blood pressure compared to normotensive patients (Zhou et al., 2021). Older age was independently associated with larger peak BP increases, with each additional decade predicting approximately 4 mmHg greater peak SBP rise. This is biologically plausible given age-related vascular stiffening and blunted baroreceptor reflexes and may warrant consideration in risk stratification for older participants.

Current ACC/AHA terminology distinguishes asymptomatic markedly elevated blood pressure, termed severe hypertension, (>180/120 mmHg without evidence of acute end-organ damage) from hypertensive emergency (the same threshold with evidence of new or worsening target-organ damage) (Jones et al., 2025). The former does not mandate urgent pharmacologic intervention in the outpatient setting. Only 2 sessions (0.4% [95% CI 0.0–1.3]) in our dataset met these criteria for ‘severe hypertension’, with no participants demonstrating clinical evidence of target-organ damage (e.g. chest pain, severe headache, focal neurologic deficits, or related visual disturbance).

The observed hemodynamic profile of psilocybin compares favorably to scenarios routinely encountered in clinical practice. Per the 2024 ACC/AHA guidelines for perioperative cardiovascular management, “there is little evidence for increased risk of perioperative complications in patients with preoperative BP <180/110 mm Hg” in the absence of ischemic heart disease, heart failure, cerebrovascular disease, insulin-treated diabetes, or creatinine >2 mg/dL (Thompson et al., 2024). Psilocybin sessions involve far less physiological stress than surgery, yet current criteria are far more restrictive.

The commonly employed exclusion criterion of blood pressure >140/90 mmHg for psilocybin clinical trials was established conservatively, in the absence of safety data. At that time, the authors of the initial guidelines for psychedelic research appropriately acknowledged “Modification of these limits may be considered in future studies if safety continues to be observed under these parameters,” (Johnson et al., 2008). We consider the findings presented here as necessary and sufficient to modify current blood pressure threshold criteria in clinical settings. Our dataset provides no evidence that a blood pressure of 140/90 is the appropriate safety boundary for excluding patients from psilocybin trials, as 6.1% sessions in this study involved patients with BP >140/90 and none experienced an adverse cardiovascular event apart from transient, asymptomatic elevations in blood pressure.

The consequences of overly restrictive cardiovascular eligibility criteria extend beyond individual potential trial participants. For example, hypertension is disproportionately common among Black adults (Ogunniyi et al., 2021), individuals with lower socioeconomic status (Xu et al., 2022), and populations with high rates of depression (Fang et al., 2022), substance use disorders (Lindenfeld et al., 2024), and PTSD (Kibler et al., 2009), some of the very indications for which psilocybin is being examined for therapeutic potential. In our ongoing trial of psilocybin for opioid use disorder (NCT06067737), 8 of 63 screened participants (12.7%) were excluded solely for hypertension despite being otherwise eligible and motivated to participate. Many individuals with substance use disorders have limited access to primary care and cannot readily optimize their blood pressure to meet stringent enrollment criteria. Excluding them based on criteria lacking empirical justification raises concerns about equity and access and does not appear congruent with the risk-benefit profile of psilocybin, which may hold therapeutic potential for OUD. Cardiovascular eligibility thresholds for psilocybin trials established without empirical justification are not neutral. Our data provide the first robust empirical basis for a rational, evidence-based revision of these criteria that could meaningfully expand access to psilocybin trials without compromising participant safety.

### Proposed criteria

Based on these data, we propose broadening eligibility to resting blood pressure <160/100 mmHg. Individuals on stable antihypertensive regimens (≥4 weeks) should be permitted if there is no history of hypertensive emergency, target-organ damage, or major cardiovascular disease (defined as prior myocardial infarction, decompensated heart failure, significant valvular disease, or unstable arrhythmia). Heart rate alone should not be an exclusion criterion. There is no clear reason to exclude based on heart rate. Exclusion of major cardiovascular disease should continue.

On dosing days, we recommend that sessions should proceed if pre-administration blood pressure is<160/100 mmHg. Physician discretion may allow dosing with baseline SBP 160–170 or DBP 100–105 mmHg. Until more empirical data are available in this small subset of patients, we recommend against psilocybin administration if SBP >170 or DBP >105 mmHg pre-dosing.

Tachycardia should not necessarily preclude psilocybin administration, since anticipatory anxiety and psychological arousal are expected contributors to elevated heart rate prior to dosing sessions. Facilitators should distinguish anxiety-driven tachycardia from autonomic tachycardia, which may require further evaluation. Participants with established supraventricular tachyarrhythmias (e.g., atrial fibrillation, Wolff-Parkinson-White), should not be enrolled.

### Monitoring protocol

We recommend blood pressure measurement at baseline, 1.5 hours, 3 hours and 6 hours. This schedule captures the peak psilocybin hemodynamic response window and the resolution phase, while avoiding excessive procedural burden for monitors and patients during the psychedelic experience. If SBP ≥180 or DBP ≥110 mmHg, monitoring frequency should increase to every 30 minutes until below threshold. If elevation persists >30 minutes, the study physician should be notified; oral antihypertensives (e.g., captopril 25 mg) may be administered at physician discretion but should not be routinely used due to risks associated with rapid lowering of blood pressure. Emergency department transfer is indicated for symptoms of target-organ damage and should be considered for BP ≥200/120 mmHg refractory to on-site treatment measures. Tachycardia in an otherwise asymptomatic, cardiovascularly healthy individual does not warrant more frequent monitoring nor treatment.

### Limitations

This analysis has several limitations. All data came from a single site (Johns Hopkins), and encompass a racially homogeneous group of largely White individuals, limiting generalizability. Participants were screened populations and none had major cardiovascular disease; these findings cannot be extrapolated to individuals with cardiovascular disease.

Furthermore, the subgroup with baseline BP ≥140/90 mmHg was small (n=34 sessions, 6.1%), limiting statistical power to characterize the response distribution in this group with precision. Finally, this was a retrospective pooled analysis with heterogeneous inclusion criteria across studies. Participants were pre-screened under study-specific cardiovascular thresholds, meaning our dataset likely under-represents individuals with elevated baseline blood pressures who might have been enrolled if more permissive criteria were used.

Of note, the Johns Hopkins Medicine Institutional Review Board conducted an audit of the JHU site of Griffiths et al. (2025) and required disclosure of the following in all related publications: (1) two unapproved study team members, one of whom was also a funding sponsor, were directly engaged in the research; (2) an approved study team member who led the qualitative analysis was also a funding sponsor, which was not disclosed to the IRB; (3) conflicts of interest related to these individuals were not appropriately disclosed or managed; and (4) funding sponsorship was not disclosed to the IRB. These issues have not otherwise impacted the analyses presented here.

## Conclusion

In 536 dosing sessions, psilocybin produced predictable, transient peak blood pressure increases equivalent to moderate exercise or sex, and heart rate increases were even milder. Periods of elevated blood pressure typically lasted less than 15 minutes. Current baseline blood pressure exclusion criteria of >140/90 mmHg are not supported by these data and may unnecessarily restrict access to psilocybin therapy for populations with the most to gain from its potential therapeutic effects. We propose broadening blood pressure eligibility criterion to <160/100 mmHg (with dosing-day flexibility up to 170/105 mmHg at physician discretion), permitting stable antihypertensive use, while maintaining exclusions for established cardiovascular disease. Heart rate elevations were modest and self-limited; there is no basis for HR-based exclusion. Six hours of monitoring appears sufficient for uncomplicated sessions, with escalation protocols in place for the small number of dosing sessions involving sustained higher blood pressure elevation. Prospective enrollment of participants with baseline BP between 140/90 and 160/100 mmHg is both a logical next step in this line of research and an ethical imperative given the access implications of continued overly conservative cardiovascular exclusion criteria.

## Data Availability

De-identified data from this pooled analysis are available from the corresponding author upon reasonable request, subject to a data use agreement and institutional review board approval at the requesting institution. Data from individual studies remain subject to the data sharing policies of the original study protocols.

## Sources of Funding

The Center for Psychedelic and Consciousness Research is supported by Tim Ferriss, Matt Mullenweg, Craig Nerenberg, Blake Mycoskie, and the Steven and Alexandra Cohen Foundation. SMN, DBY, and BW have received funding from the Gracias Family Foundation. SMN and BW have received funding from Reason for Hope and Fifth Generation. BSB receives research support from Abbott Laboratories, Compass Pathways, Definium Therapeutics, and Reunion Neuroscience.

## Disclosures

SMN has consulted for Resilient Pharmaceuticals, and MMS Holdings. BW owns Axial Therapeutic Research, a company investigating the safety and effectiveness of alternative treatments for military veteran health. DBY has personally received a consulting fee from Soneira and speaking fees from Integrative Psychiatry Institute. AGR is a paid scientific advisor to Otsuka Pharmaceutical Development & Commercialization Inc., and the Chair of the Scientific Advisory Board for Psyence BioMed. BSB holds stock options for CB Therapeutics and has previously held stock in AtaiBeckley Inc and GH Research in the past three years. In the past three years he has served or currently serves as an advisor for AbbVie, CB Therapeutics, GH Research, Livanova, Janssen Pharmaceuticals (Johnson & Johnson), and MindMed (Definium Therapeutics). He also receives monetary compensation for editorial work from DynaMed Plus (EBSCO Industries, Inc) and speaker’s fees from Toronto-Dominion Bank. FSB has consulted for MindState Design Labs LLC, Wavepaths Ltd, and Gilgamesh Pharmaceuticals Inc.

## Notes

### Author Declarations

The Johns Hopkins Medicine IRB approved this analysis (IRB00548489) via expedited review.

### Summary of Updates

Fixed error in demographic calculations. Updated author disclosures. Otherwise minor textual edits.

