## Supplement for "Acute Cardiovascular Effects of Psilocybin: A Pooled Analysis of 14 Studies with Safety Recommendations"

### Supplementary Materials

#### Supplementary Methods: Mixed-Effects Models

To assess whether psilocybin dose or session number predicted hemodynamic response, we fit linear mixed-effects models with change from baseline in systolic and diastolic blood pressure as outcomes. Models included natural splines (4 degrees of freedom) for time since dose to capture the nonlinear time course, fixed effects for dose (mg) and session number, and random intercepts for participant and study to account for repeated measures and clustering. Models were fit using restricted maximum likelihood (REML) in R version 4.4.1 (lme4 package).

**Model specification:**

```
    library(lme4)
    library(splines)


m_sbp <- lmer(sbp_delta ~ ns(time_since_dose, df = 4) + dose + session +
                (1 | study/vol_id),
              data = d_model)


m_dbp <- lmer(dbp_delta ~ ns(time_since_dose, df = 4) + dose + session +
                (1 | study/vol_id),
              data = d_model)
```

#### Supplementary Results

Dose was not significantly associated with peak change in SBP ($\beta$ = 0.07 mmHg per mg, SE = 0.05, p = 0.16) or DBP ($\beta$ = 0.07 mmHg per mg, SE = 0.04, p = 0.05). Session number did not predict SBP ($\beta$ = -0.18, SE = 0.37, p = 0.62) or DBP ($\beta$ = -0.19, SE = 0.26, p = 0.46) response.

Individual-level variance accounted for 32% (SBP) and 29% (DBP) of total variance, whereas study-level variance accounted for 7% and 4%, respectively.

**Supplementary Table 1. Linear mixed-effects model for systolic blood pressure change from baseline**

| Fixed Effects | Estimate | SE | z-value | p-value |
|---|---|---|---|---|
| Intercept | -6.66 | 3.43 | -1.94 | 0.05 |
| Time (spline 1) | 23.71 | 3.03 | 7.83 | <0.001 |
| Time (spline 2) | 1.48 | 2.09 | 0.71 | 0.48 |
| Time (spline 3) | 3.68 | 6.22 | 0.59 | 0.55 |
| Time (spline 4) | 14.48 | 3.83 | 3.78 | <0.001 |
| Dose (per mg) | 0.07 | 0.05 | 1.39 | 0.16 |
| Session | -0.18 | 0.37 | -0.50 | 0.62 |

| Random Effects | Variance | SD | % Total |
|---|---|---|---|
| Participant (intercept) | 62.85 | 7.93 | 32% |
| Study (intercept) | 14.70 | 3.83 | 7% |
| Residual | 119.79 | 10.95 | 61% |

N = 5,268 observations; 368 participants; 14 studies

**Supplementary Table 2. Linear mixed-effects model for diastolic blood pressure change from baseline**

| Fixed Effects | Estimate | SE | z-value | p-value |
|---|---|---|---|---|
| Intercept | -5.16 | 2.47 | -2.09 | 0.04 |
| Time (spline 1) | 14.13 | 2.24 | 6.31 | <0.001 |
| Time (spline 2) | -1.52 | 1.55 | -0.98 | 0.33 |
| Time (spline 3) | 5.17 | 4.59 | 1.13 | 0.26 |
| Time (spline 4) | 11.83 | 2.83 | 4.18 | <0.001 |
| Dose (per mg) | 0.07 | 0.04 | 1.96 | 0.05 |
| Session | -0.19 | 0.26 | -0.74 | 0.46 |

| Random Effects | Variance | SD | % Total |
|---|---|---|---|
| Participant (intercept) | 28.08 | 5.30 | 29% |
| Study (intercept) | 3.90 | 1.98 | 4% |
| Residual | 65.67 | 8.10 | 67% |

N = 5,269 observations; 368 participants; 14 studies
